# Impact of BNT162b2 mRNA Vaccination on the Development of Short and Long-term Vaccine-Related Adverse Events in Inflammatory Bowel Disease: A Multi-Center Prospective Study

**DOI:** 10.1101/2022.02.22.22271342

**Authors:** Mohammad Shehab, Fatema Alrashed, Israa Abdullah, Ahmad Alfadhli, Hamad Ali, Mohamed Abu-Farha, Arshad Channanath, Jehad Abubaker, Fahd Al-Mulla

## Abstract

**Introduction:** SARS-CoV-2 vaccination has been effective in protecting against severe COVID-19 infections and related mortality. It is recommended for all individuals including patients with inflammatory bowel disease (IBD). However, safety data is lacking in this group of patients. Therefore, we aim to evaluate the short- and long-term vaccine related adverse events (AEs) in patients with IBD.

**Method:** This is a prospective, observational cohort study investigating short- and long-term AEs related to BNT162b2 vaccine in patients with IBD (study group) after first and second dose compared to healthy participants (study group). Patients were recruited at the time of attendance to clinic or infusion rooms. Short term (<3 weeks) localized and systemic AEs were assessed via questionnaire. Follow-up phone-based survey was made to collect data on long term (up to 24 weeks) AEs.

**Results:** A total of 408 patients answered the questionnaires, 204 patients in each group, the study and control group. No serious adverse events were reported in either the study nor the control group after the first or the second dose. Participants in the control group reported more frequent pain at the injection site than those in the study group after the first dose (58 (57%) vs 38 (37%) respectively, P-value= 0.005). After the second dose, tiredness was reported more frequently in the control group [49 (48%)] compared to the study group [25 (24%), (P-value<0.001)]. At 20-24 weeks post vaccination, 386 out of 408 (94.6%) patients were willing to participate in the follow-up phone based questionnaire (196 (96.1%) in the study group vs 190 (93.1%) in the control group). In both groups, none of the patients reported local, systemic or severe adverse events (0 out of 386) at week 20-244 post second dose.

**Conclusion:** The BNT162b2 vaccine is safe in patients with IBD. No severe or long-term adverse events were reported in our study. The frequency of local and systemic adverse events after the second dose was generally higher among healthy participants compared to patients with IBD. Further studies including a larger cohort with longer follow-up duration are needed to assess for possible rare adverse events.

## Introduction

The severe acute respiratory syndrome coronavirus 2 (SARS-CoV-2) emerged in Wuhan, China in December 2019.^1^ The disease has been known to cause a significant morbidity and mortality among many of those infected. The outbreak, which was later declared a pandemic, had global health and socioeconomic consequences.^2^ This has led to an international effort for vaccine development and the introduction of the first vaccine, the BNT162b2 mRNA (Pfizer/BioNTech) vaccine in December 2020 followed by ChAdOx1 nCoV-19, mRNA-1273 (Moderna) and other vaccines, which later were authorized under the emergency use authorization.^3,4^

Thereafter, clinical trials and real-world data have shown efficacy and safety of these vaccines in reducing COVID-19 infection severity and decreasing both hospitalization and mortality in patients with COVID-19. Nevertheless, patients with inflammatory bowel disease (IBD) were largely excluded from these trials.^5–7^

Despite the fact that many of the patients with IBD are on immune-modifying medications, those patients were not found to be at higher risk of developing COVID-19 infection. However, being on corticosteroids was found to be a risk factor for developing more severe infection.^8^ Vaccination against the SARS-CoV-2 virus in IBD patients is highly recommended by most international gastrointestinal societies.^9,10^

A study showed that the overall prevalence of COVID-19 vaccination among patients with IBD on biologic therapies was lower than that of the general population.^11^ Furthermore, many studies focused on the efficacy of COVID-19 vaccination in patients with IBD receiving biologic therapies, while the safety of vaccination was not extensively explored.^12,13^ However, evidence regarding the vaccine safety in IBD patients is slowly emerging, with majority of studies investigating only the short-term adverse events following vaccination in patients with IBD.^14,15^ Therefore, it is imperative to assess the long-term safety of SARS-CoV-2 vaccine in patients with IBD. This study aims to evaluate the short-and long-term adverse events following vaccination with BNT162b2 mRNA vaccine among patients with IBD.

## Material and Methods

We performed a prospective multi-center cohort study at two tertiary care centers (Muabark Alkabeer Hospital and Dasman Center) to assess short and long-term adverse events related to COVID-19 mRNA vaccine, BNT162b2 (Pfizer/BioNTech) in patients with IBD (study group) compared to healthy participants (control group).

This study was performed and reported in accordance with Strengthening the Reporting of Observational Studies in Epidemiology (STROBE) guidelines.^16^ This study was reviewed and approved by the Ethical Review Board of Mubarak Alkabeer Hospital and Dasman Center “Protocol # RA HM-2021-008” as per the updated guidelines of the Declaration of Helsinki (64th WMA General Assembly, Fortaleza, Brazil, October 2013) and of the US Federal Policy for the Protection of Human Subjects. The study was also approved by the regional health authority (reference: 3799, protocol number 1729/2021). Subsequently, patient informed written consent was obtained before inclusion in the study.

Localized and systemic adverse events to BNT162b2 vaccine were assessed via paper questionnaires at the time of attendance at the gastroenterology infusion rooms and outpatient clinics from August 1st, 2021, to September 15th, 2021, and patients were followed up to February 10th, 2022, using a phone based questionnaire. Outcomes were stratified by first and second dose.

Study group patients were eligible to be included if they: (1) had confirmed diagnosis of inflammatory bowel disease (IBD) before the start of the study, (2) had received one or two doses of COVID-19 vaccination with BNT162b2 (Pfizer-BioNTech), (3) were at least 18 years of age or older. Patients were excluded if they received any vaccine other than the BNT162b2 or if they tested positive SARS-CoV-2 previously or had symptoms of COVID-19 since the start of the pandemic up to the time of vaccination. Patients were also excluded if they have one of the following within 8 weeks of vaccination: stool fecal calprotectin levels >250 ug/g, C-Reactive Protein (CRP) levels >10 mg/ml, active symptoms of IBD or endoscopic active disease (see below), use of corticosteroids active extraintestinal manifestation of IBD (e.g. inflammatory uveitis, arthritis, skin rashes, etc).

Patients with Harvey Bradshaw Index (HBI) >4 and partial clinical Mayo score >1 are considered to have active symptoms of IBD. In addition, patients who had colonoscopies with an endoscopic Mayo score >1 for ulcerative colitis or Simple Endoscopic Score for Crohn’s Disease (SES-CD) >4 are considered to have active endoscopic. In addition, patients who had severe allergic reaction to a previous vaccine in their life or were unwilling to participate in the study were excluded.

In the other hand, Healthy participants (control) group were individuals who volunteered to participate for the study at Dasman Center with no previous history of chronic medical illnesses such as diabetes, hypertension, cardiovascular disease, autoimmune diseases, osteoarthritis, chronic obstructive pulmonary disease, renal disease, asthma, hyperlipidemia, or history of stroke and bleeding disorder. In addition, basic laboratory tests were performed (full blood count, renal function tests, liver function tests, lipid profile, HbA1c, ESR, and CRP) to objectively screen for underlying diseases.

Baseline questionnaire assessed type of immunization, date of immunization(s), patient demographics and IBD characteristics, and data regarding IBD medication use around the time of vaccination. Participants were asked to report short-term localized and systemic adverse events defined as adverse events occurring within 14 days after receiving BNT162b2 vaccine dose 1 and 21 days of receiving vaccine dose 2. The follow-up phone-based survey collected data on long term (20-24 weeks from first dose) adverse events of BNT162b2 vaccination.

Vaccine adverse events were classified as injection site (localized) or systemic reactions. Adverse localized reactions included pain, redness, itching, swelling, or tenderness at the injection site. Systemic adverse reactions included fever, chills, fatigue, headache, joint pain, muscle aches, nausea, allergic reaction, rash, or other. Severe adverse events were defined as incidence of pulmonary embolism, acute myocardial infarction, immune thrombocytopenia, and disseminated intravascular coagulation per the Food and Drug Administration (FDA) definition of potential adverse events of interest.^17^

Diagnosis of inflammatory bowel disease (IBD) was made according to the international classification of diseases (ICD-10 version:2016). Patients were considered to have IBD when they had ICD-10 K50, K50.1, K50.8, K50.9 corresponding to Crohn’s disease (CD) and ICD-10 K51, k51.0, k51.2, k51.3, k51.5, k51.8, k51.9 corresponding to ulcerative colitis (UC).^18^

### Statistical Analysis

Analysis was conducted using R (R Core Team, 2017). We performed descriptive statistics to present the demographic characteristics of patients included in this study. McNemar test was used to determine whether the proportion of participants who had any symptoms (yes or no) after the first dose of vaccine differed after the second dose. Pearson’s Chi-squared test was used to compare the proportions of symptoms of the control group vs. study group. Participants in both groups were matched for *Age* and *Gender*. The technique attempts to choose matches that collectively optimize an overall criterion. The criterion used is the sum of the absolute pair distances in the matched sample.

## Results

### Baseline characteristics

Between August 1st 2021 and September 15th, a total of 204 patients diagnosed with inflammatory bowel disease (IBD) answered the questionnaire. Of these, 119 (58%) were males. Median age of the patients included is 34.6 years (IQR 25-41). 140 (68.7%) patients and 64 (31.3%) patients had Crohn’s disease and ulcerative colitis, respectively. Most patients were receiving biologic therapy [82(40%)], followed by immunomodulators [75 (37.0%)]. Whereas 47 (23.0%) patients were on 5-aminosalicylates.

In the study group, half of the patients (n=102) received one dose of Pfizer-BioNTech vaccine, while the other half (n=102) received two doses of the vaccine. Asthma (8%), arthritis (5%), and diabetes (3.9%) were the most common comorbidities in the study group. Demographics are shown in table 1. No serious adverse events were reported in either the study nor the control group after the first or the second dose.

**Table 1:** Baseline Characteristics of Patients with Inflammatory Bowel Disease (IBD)

| Characteristic | N = 204 |
| --- | --- |
| **Age (years)<sup>1</sup>** | 34.6 (25, 41) |
| **Gender** |  |
| Female | 85 (42%) |
| Male | 119 (58%) |
| **Nationality** |  |
| Kuwaiti | 180 (88%) |
| Non-Kuwaiti | 24 (12%) |

| Disease extent, n (%) |  |
| --- | --- |
| **Ulcerative colitis (UC)** | 64 (31.3%) |
| E1: ulcerative proctitis | 12 (18.7%) |
| E2: left sided colitis | 20 (31.2%) |
| E3: extensive colitis | 32 (50.0%) |
| **Crohn's disease (CD)** | 140 (68.7%) |
| L1: ileal | 62 (44.2%) |
| L2: colonic | 24 (17.1%) |
| L3: ileocolonic | 48 (34.4%) |
| L4: upper gastrointestinal | 6 (4.3%) |
| B1: inflammatory | 68 (48.8%) |
| B2: stricturing | 33 (23.3%) |
| B3: penetrating | 39 (27.7%) |
| **Past Medical History** |  |
| Diabetes | 8 (3.9%) |
| Hypertension | 5 (2.0%) |
| Heart Disease | 5 (2.0%) |
| Arthritis | 11 (5.0%) |
| COPD | 2 (1.0%) |
| Kidney | 5 (2.0%) |
| Asthma | 16 (8.0%) |
| Hyperlipidemia | 7 (3.0%) |
| **IBD Medications** |  |
| 5- aminosalicylates | 47 (23.0%) |
| Immunomodulators | 75 (37.0%) |
| Biologics and small molecules | 82 (40.0%) |

<sup>1</sup>Median (IQR) or Frequency (%)

### Symptoms after the first dose in the study group vs control group

Adverse events after the first vaccine dose are shown in table 2. The most common local adverse event was pain at the injection site reported in 37% of the study group. In general, after receiving the first dose, local adverse events were reported more frequently by patients in the control group. Specifically, subjects in the control group reported more frequent pain at the injection site than those in the study group (58 (57%) vs 38 (37%) respectively, P-value= 0.005). Redness and swelling at the injection site were also more common in the control group compared to the study group, 17 (17%) vs 3 (2.9%), P-value <0.001, and 18 (18%) vs 4 (3.9%), P-value=0.002, respectively.

**Table 2:** Comparison of symptoms after First Dose in patients with IBD (study) group vs. healthy participants (control) group.

| Characteristic | Study Group, N = 102<sup>1</sup> | Control Group, N = 102<sup>1</sup> | p-value<sup>2</sup> |
| --- | --- | --- | --- |
| Age_in years | 33 (26, 41) | 34 (28, 43) |  |
| Gender |  |  |  |
| Female | 48 (47%) | 52 (51%) |  |
| Male | 54 (53%) | 50 (49%) |  |
| Injection Site |  |  |  |
| Pain | 38 (37%) | 58 (57%) | 0.005 |
| Redness | 3 (2.9%) | 17 (17%) | <0.001 |
| Swelling | 4 (3.9%) | 18 (18%) | 0.002 |
| Systemic AEs |  |  |  |
| Tiredness | 19 (19%) | 35 (34%) | 0.011 |
| Headache | 15 (15%) | 16 (16%) | 0.8 |
| Muscle pain | 11 (11%) | 24 (24%) | 0.016 |
| Chills | 4 (3.9%) | 14 (14%) | 0.014 |
| Fever | 13 (13%) | 14 (14%) | 0.8 |
| Nausea | 1 (1.0%) | 9 (8.8%) | 0.009 |
| Joint pain | 8 (7.8%) | 11 (11%) | 0.5 |

<sup>1</sup>Median (IQR) or Frequency (%)
<sup>2</sup>Wilcoxon rank sum test; Pearson's Chi-squared test

Similarly, after receiving the first dose, more subjects reported systemic reactions in the control group compared to the study group. Specifically, tiredness was reported by 35 (34%) of the subjects in the control group as opposed to 19 (19%) subjects in the study group (P-value= 0.011), muscle pain in 24 (24%) subjects in the control group and 11 (11%) subjects in the study group (P-value=0.016), chills in 14 (14%) subjects in the control group compared to 4 (3.9%) subjects in the study group (P-value=0.014), and nausea in 9 (8.8%) subjects from the control group compared to 1 (1.0%) subjects in the study group (P-value=0.009). There was no significant difference in the occurrence of headaches, fever or joint pain between the control group and the study group after the first dose of the vaccine.

### Symptoms after the second dose in the study group vs control group

The frequency of local adverse events after the second dose was also generally higher among subjects in the control group than those in the study group (table 3). Pain at the injection site was reported by 50 (49%) subjects in the control group and 32 (31%) subjects in the study group (P-value=0.01). Additionally, 15 (15%) subjects in the control group reported redness at the injection site compared to 4 (3.9%) subjects in the study group (P-value=0.008), while 29 (28%) subjects had swelling at the injection site in the control group compared to 3 (2.9%) subjects in the study group (P-value <0.001).

**Table 3:** Comparison of symptoms after Second Dose -in patients with IBD (study) group vs. healthy participants (control) group.

| Characteristic | Study Group, N = 102<sup>1</sup> | Control Group, N = 102<sup>1</sup> | p-value<sup>2</sup> |
| --- | --- | --- | --- |
| Age_in years | 34 (25, 41) | 33 (25, 45) |  |
| Gender |  |  |  |
| Female | 43 (42%) | 49 (48%) |  |
| Male | 59 (58%) | 53 (52%) |  |
| Injection Site |  |  |  |
| Pain | 32 (31%) | 50 (49%) | 0.010 |
| Redness | 4 (3.9%) | 15 (15%) | 0.008 |
| Swelling | 3 (2.9%) | 29 (28%) | <0.001 |
| **Systemic AEs** |  |  |  |
| Tiredness | 25 (24%) | 49 (48%) | <0.001 |
| Headache | 21 (20%) | 35 (34%) | 0.001 |
| Muscle pain | 16 (15%) | 34 (33%) | <0.001 |
| Chills | 9 (8.8%) | 19 (19%) | 0.042 |
| Fever | 20 (20%) | 26 (25%) | 0.3 |
| Nausea | 2 (2.0%) | 14 (14%) | 0.002 |
| JointPain | 4 (3.9%) | 20 (20%) | <0.001 |

<sup>1</sup>Median (IQR) or Frequency (%)
<sup>2</sup>Wilcoxon rank sum test; Pearson's Chi-squared test

When comparing systemic reactions among subjects in the control group as opposed to the study group, tiredness was reported by 49 (48%) subjects vs 25 (24%) subjects (P-value<0.001), headaches in 35 (34%) subjects vs 21 (20%) (P-value=0.001), muscle pain in 34 (33%) vs 16 (15%) subjects (P-value <0.001), chills in 19 (19%) vs 9 (8.8%) subjects (P-value=0.042), nausea in 14 (14%) subjects vs 2 (2.0%) subjects (P-value=0.002), and joint pain in 20 (20%) vs 4 (3.9%) subjects (P-value<0.001) respectively. Conversely, no significant difference was found in the frequency of fever between the control group and the study group (26 (25%) vs 20 (20%), P-value= 0.3). (table 3).

Other than nausea, none of the participants reported gastrointestinal (GI) related symptoms such as diarrhea or abdominal pain after the first or second dose. Furthermore, none of the patients reported any severe adverse events after the first or second dose. Additionally, no significant differences in any adverse reaction frequency were seen based on sex, or age.

### Symptoms after the first dose vs the second dose in the study group

The frequency and type of adverse reactions after the first dose of the vaccine was compared with adverse reactions after the second dose of the vaccine among patients with IBD (study group). There was no significant difference in the frequency of symptoms reported after the first and second dose among subjects in the study group. The most common local reaction was pain at the injection site reported by 38 (37%) patients after the first dose and 32 (31%) patients after the second dose (P-value= 0.3). Redness was reported in 3 (2.9%) subjects after the first dose and in 4 (3.9%) after the second dose (P-value>9) and swelling in 4 (3.9%) subjects after the first dose and in 3 (2.9%) after the second dose (P-value>9).

Systemic reactions were reported as follows: tiredness in 19 (19%) after dose 1 vs 25 (24%) after dose 2, P-value >9, headache in 15 (15%) after dose 1 vs 21 (20%) after dose 2, P-value >9, muscle pain in 11 (11% after dose 1 vs 16 (15%) after dose 2, P-value >9, chills in 4 (3.9%) after dose 1 vs 9 (8.8%) after dose 2, P-value= 0.2, fever in 13 (13%) after dose 1 vs 20 (20%) after dose 2, P-value= 0.2, nausea in 1 (1%) after dose 1 vs 2 (2%) after dose 2, P-value >9, and joint pain in 8 (7.8%) after dose 1 vs 4 (3.9%) after dose 2, P-value= 0.3 (table 4).

**Table 4:**
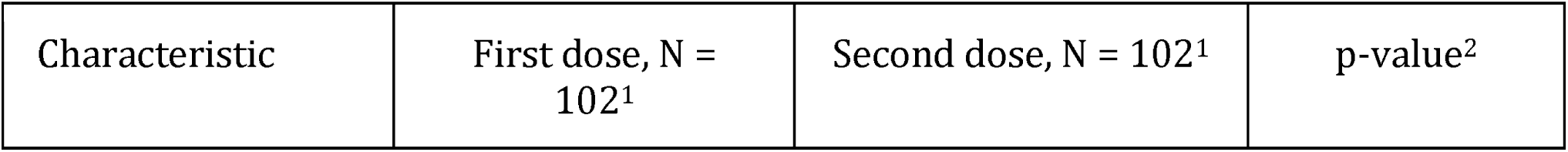
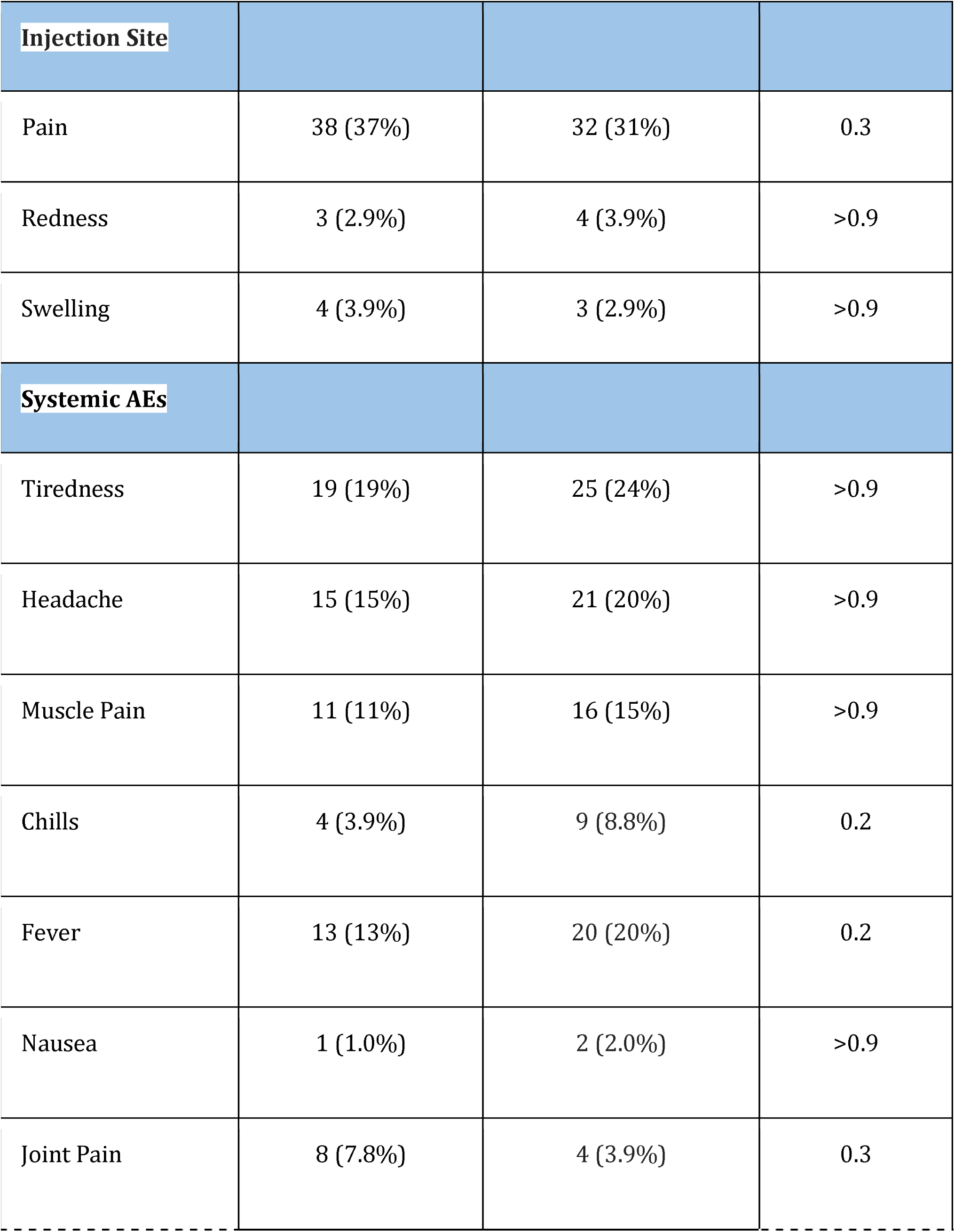
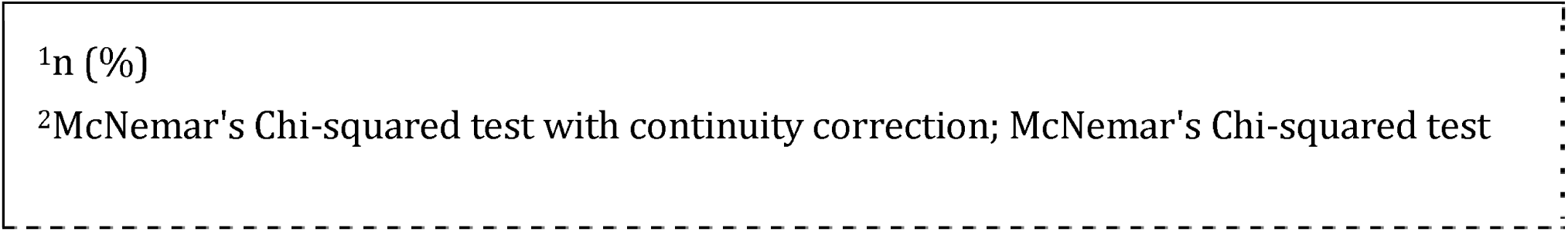
Symptoms after first dose and second dose in patients with IBD (study group)

| Characteristic | First dose, N = 102<sup>1</sup> | Second dose, N = 102<sup>1</sup> | p-value<sup>2</sup> |
| --- | --- | --- | --- |

| Injection Site |  |  |  |
| --- | --- | --- | --- |
| Pain | 38 (37%) | 32 (31%) | 0.3 |
| Redness | 3 (2.9%) | 4 (3.9%) | >0.9 |
| Swelling | 4 (3.9%) | 3 (2.9%) | >0.9 |
| Systemic AEs |  |  |  |
| Tiredness | 19 (19%) | 25 (24%) | >0.9 |
| Headache | 15 (15%) | 21 (20%) | >0.9 |
| Muscle Pain | 11 (11%) | 16 (15%) | >0.9 |
| Chills | 4 (3.9%) | 9 (8.8%) | 0.2 |
| Fever | 13 (13%) | 20 (20%) | 0.2 |
| Nausea | 1 (1.0%) | 2 (2.0%) | >0.9 |
| Joint Pain | 8 (7.8%) | 4 (3.9%) | 0.3 |

<sup>1</sup>n (%)
<sup>2</sup>McNemar's Chi-squared test with continuity correction; McNemar's Chi-squared test

### *Long-term* adverse events

At 20-24 weeks post vaccination, 386 out of 408 patients were willing to participate in the follow-up phone based questionnaire. In the study group, 196 (96.1%) patients and 190 (93.1%) in the control group answered the questionnaire. 21 (10.7%) out of 196 patients in the study group, and 19 (10.0%) out of 190 in the control group reported to have breakthrough SARS-CoV-2 infection confirmed by PCR test after second dose of vaccination. None of the patients who tested positive were hospitalized.

In both groups, none of the patients reported local, systemic, or severe adverse events (0 out of 386). In addition, short-term local and systemic adverse events have been resolved in the control and study group.

## Discussion

We performed a survey based study to explore the onset of adverse events related to the BNT162b2 vaccine in patients with inflammatory bowel disease (IBD) compared with healthy participants. In our study, none of our patients had severe vaccine-related adverse events, as they are very rare.^19^ We found that the most common adverse events after the first and second dose were tiredness and headache, followed by local pain at the injection site. Nausea was reported in both study and control group, however, none of the group reported other gastrointestinal (GI) related symptoms such as diarrhea or abdominal pain.

Weaver et al^20^ explored vaccine related adverse events among patients with IBD and the effect of vaccination on IBD disease course. Similar to our study, they found that Severe localized and systemic vaccine-related adverse events were rare in patients with IBD. Injection site tenderness (68%) and fatigue (46% dose 1, 68% dose 2) were the most commonly reported localized and systemic adverse events after vaccination.

Interestingly, we also found that local and systemic adverse events were more common in healthy participants compared to patients with IBD. Given that the majority of our IBD cohort are on biologics or immunomodulators, it is possible that these medications blunt the immune response to vaccination. Botwin et al^14^ evaluated post-mRNA vaccination adverse events in 246 vaccinated adults with IBD participating in a longitudinal vaccine registry. Similar to our finding, the study found that adverse events were less common in individuals receiving biologic therapy. The authors concluded that patients with IBD can be reassured that the risk of adverse events is likely not increased, and may be reduced, while receiving concomitant biologic therapy.

None of the participants in our study experienced severe short-or long-term adverse events. One systematic review and meta-analysis evaluated SARSCoV-2 vaccination in IBD patients. The study did not find any severe adverse events or vaccine-related mortality in patients with IBD and that the majority of patients reported mild adverse events after vaccination, including fatigue, headache, dizziness, and gastrointestinal symptoms.^21^

Another study^22^ observed a higher rate of diarrhea and abdominal pain in vaccinated patients with IBD compared to the general population. They also found that age and disease remission was inversely correlated with onset of GI symptoms. To our knowledge, no evidence has emerged of IBD flare-ups caused by COVID-19 vaccination. Furthermore, in a population-based study, the effect of SARS-CoV-2 vaccination on IBD course was evaluated for a period of 4 weeks in patients with IBD. The study reported no clinical and laboratory exacerbation compared with pre-vaccination baseline no increase in corticosteroid prescription 1 month after vaccination in a large retrospective cohort compared with a matched unvaccinated cohort.^23^

Another study^15^ explored immediate (within one day) adverse events after mRNA SARS-CoV-2 vaccines in patients with IBD in the United States. Similar to our study, the authors found that the incidence of adverse events including acute myocardial infarction, anaphylaxis, facial nerve palsy and coagulopathy in IBD patients after COVID-19 vaccination was small and similar to a matched cohort of patients without IBD. Immediate adverse events after vaccination were rare in both cohorts.

In our study, none of our patients reported long-term adverse events 20-24 weeks after vaccination. One study^19^ involved more than 1.5 million BNT162b2 vaccinated persons from an integrated healthcare organization, followed over a period of 42 days. The study reported an excess risk of lymphadenopathy (78.4 events per 100,000 persons), herpes zoster infection (15.8 events), appendicitis (5.0 events), and myocarditis (2.7 events) in the vaccinated cohort. However, the author concluded that their results indicate that SARS-CoV-2 infection is itself a very strong risk factor for myocarditis, and it also substantially increases the risk of multiple other serious adverse events.

Taken together, these emerging data provide reassurance that to COVID-19 vaccination does not cause severe adverse events in patients with IBD and support recent consensus recommendations to vaccinate all patients with IBD. British Society of Gastroenterology (BSG),^10^ the International Organization for the Study of Inflammatory Bowel Diseases (IOIBD),^24^ and the Canadian Association of Gastroenterology^9^ recommend that all patients with IBD should receive SARS-CoV-2 vaccination regardless whether patients were in remission or not.

Our study has several strengths. It provides real world data for the public about adverse effects and vaccine safety in a subpopulation that was not studied in the initial clinical trials. In addition, the comparison to healthy participants and the rigorous matching allowed for precise estimation of the rate of adverse events in patients with IBD. Finally, the long-term follow-up period helps detect any possible late events that may occur several weeks after vaccination.

Our study also has some limitations. Given the observational nature of this study, it is possible that some hidden confounding variables were still not properly addressed. In addition, this study was performed prospectively in the context of an ongoing pandemic, therefore, association between any breakthrough infection and any given adverse events cannot be ruled out. However, in our follow-up phone-based survey we asked patients regarding any previous or current SARS-CoV-2 infection. Despite these limitations, our study provides highly anticipated data regarding the short-and long-term adverse events of the SARS-CoV-2 vaccination in patients with IBD.

## Conclusion

The BNT162b2 vaccine is safe in patients with IBD. No severe or long-term adverse events were reported in our study. The frequency of local and systemic adverse events after the second dose was generally higher among healthy participants compared to patients with IBD. Further studies including a larger cohort with longer follow-up duration are needed to assess for possible rare adverse events.

## Data Availability

ll data produced in the present study are available upon reasonable request to the authors

## Declaration

All authors declare no conflict of interest.

## Acknowledgment

none

## Contribution to the field statemen

The safety of COVID-19 vaccines have been a global concern for the general public, especially patients with autoimmune diseases such as inflammatory bowel disease (IBD). In this study we looked at BNT162b2 mRNA vaccine adverse events in IBD patients compared to healthy individuals for up to 24 weeks post vaccination. We found that the vaccine was safe in both groups with minor adverse events. We also found that patients with IBD are less likely to develop vaccine related adverse events.

